# A multicenter evaluation of a novel microfluidic rapid AST assay for Gram-negative bloodstream infections

**DOI:** 10.1101/2024.03.20.24304581

**Authors:** Benjamin Berinson, Emma Davies, Jessie Torpner, Linnea Flinkfeldt, Jenny Fernberg, Amanda Åman, Johan Bergqvist, Håkan Öhrn, Jonas Ångström, Cecilia Johansson, Klara Jäder, Helena Andersson, Ehsan Ghaderi, Maria Rolf, Martin Sundqvist, Holger Rohde, Teresa Fernandez-Zafra, Christer Malmberg

## Abstract

**Objectives:** Common phenotypic methods for antibiotic susceptibility testing (AST) of bacteria are slow, labour intensive and display considerable technical variability. The QuickMIC system provides rapid AST using a microfluidic linear gradient. Here we evaluate the performance of QuickMIC at four different laboratories with regards to speed, precision, accuracy, and reproducibility in comparison to broth microdilution (BMD).

**Methods:** Spiked blood cultures (n=411) and clinical blood cultures (n=148) were tested with the QuickMIC Gram negative (GN) panel and compared with BMD for the 12 on-panel antibiotics, and 10 defined strains were run at each site to measure reproducibility. Logistic and linear regression analysis was applied to explore factors affecting assay performance.

**Results:** The overall essential agreement (EA) and categorical agreement (CA) between QuickMIC and BMD were 95.6% and 96.0%, respectively. Very major error (VME), Major error (ME) and minor error (mE) rates were 1.0, 0.6 and 2.4%, respectively. Inter-laboratory reproducibility between the sites was high at 98.9% using the acceptable standard of ±1 log2 unit. The mean in-instrument analysis time overall was 3h 13 min (SD: 29 min). Regression analysis indicated that QuickMIC is robust with regards to initial inoculate and delay time after blood culture positivity.

**Conclusions:** We conclude that QuickMIC can be used to rapidly measure MIC directly from blood cultures in clinical settings, with high reproducibility, precision, and accuracy. The microfluidics-generated linear gradient ensures high repeatability and reproducibility between laboratories, thus allowing a high level of trust in MIC values from single testing, at the cost of reduced measurement range compared to BMD.

## 1 Introduction

Antimicrobial resistance (AMR) is a global public health concern, posing significant challenges to effective infection management. The inappropriate use of antibiotics contributes to the emergence and spread of resistant pathogens, leading to increased morbidity, mortality, and healthcare costs [1]. Diagnostic interventions such as bacterial identification, syndromic testing and especially antimicrobial susceptibility testing (AST) play a critical role in guiding appropriate antibiotic therapy, especially in critical disease such as bloodstream infection (BSI) [2]. Mortality rates due to bloodstream infection range between 12% and 32% in North America and Europe and are even higher in low- and middle-income countries [3,4]. Mortality is due in part to increasing rates of antimicrobial-resistant pathogens, and patients infected with resistant pathogens are more likely to receive ineffective empiric antibiotic therapy, which is associated with poor outcomes, including death [5,6]. Conversely, treatment with overly broad antibiotics increases risk of adverse drug events and drives further development of resistance [7]. Despite advances in antimicrobial susceptibility testing, the delays associated with standard of care tests can lead to suboptimal patient outcomes, increased length of hospital stays, and prolonged exposure to broad-spectrum antibiotics. Rapid antimicrobial susceptibility testing (Rapid AST or rAST) offers the potential to overcome these challenges by providing timely and accurate results, allowing for prompt adjustment of antibiotic therapy and targeted treatment selection [4]. Newly developed tools, like the Pheno® system (Accelerate Diagnostics), or Vitek® Reveal™ (Biomérieux) offer rAST results in 7h and 5.5h, respectively. EUCAST RAST disk diffusion can deliver even faster AST results (as early as 4h), comes at a low cost and has proven to be a valuable tool [8–10], but does not provide quantitative MIC values, is fairly laborious with exact intervals and manual reading (although automation is possible [11,12]), as well as only providing breakpoints for a selection of species and antibiotic agents.

QuickMIC® is an *in vitro* diagnostic system for rapid AST directly from positive blood cultures. The technology is based on microfluidics and solid phase cytometry, providing growth-based susceptibility results in 2-4 hours [13,14]. A linear antibiotic concentration gradient is generated in a 3D agarose hydrogel containing bacteria from the patient’s blood culture sample (Fig. 1). The minimum inhibitory concentration (MIC) can be rapidly identified via automated, real-time quantification of bacterial microcolony growth. The system consists of single-use, disposable QuickMIC Cassettes with pre-filled, dried antibiotics, modular QuickMIC Instrument to process the Cassettes, and QuickMIC Analyst software for automated analysis. Instruments can be stacked for increased capacity (12 can be run by a single PC). One Instrument can analyse one patient sample against a panel of 12 antibiotics per run.

**Figure 1.**
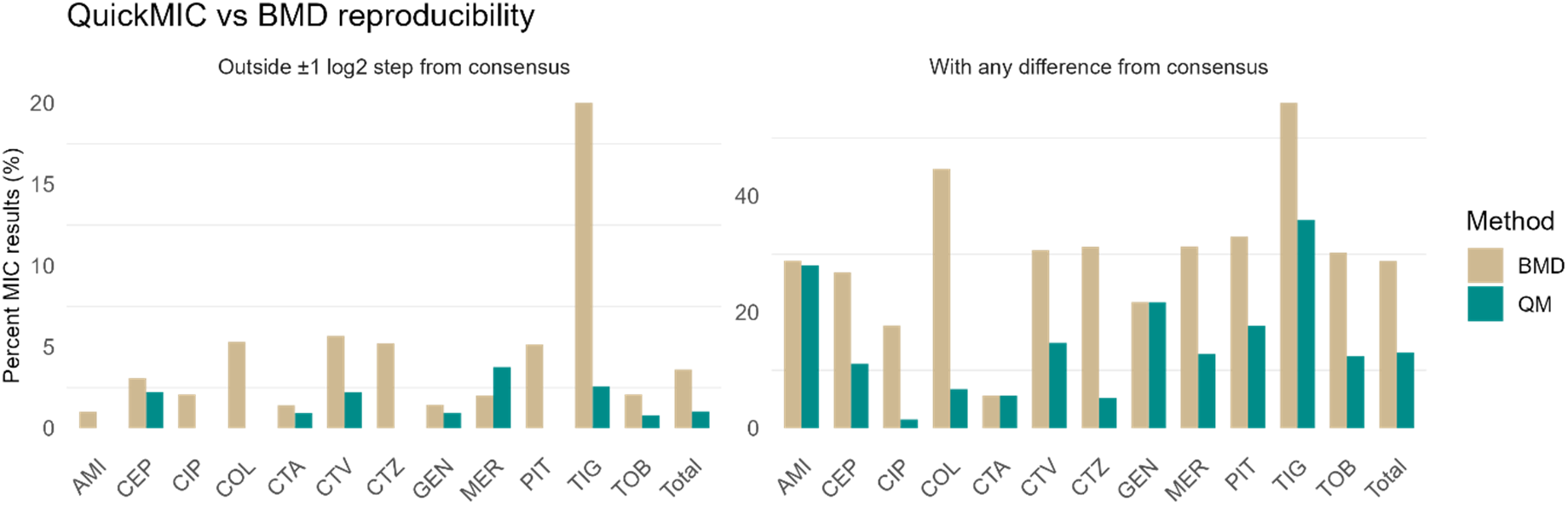
Reproducibility of QuickMIC (green) compared to BMD (brown), showing percent of results within 1 log2 step (left) for each antibiotic, and percent of all results not yielding the same MIC for each antibiotic. QuickMIC is overall more reproducible.

The aim of the current study was to evaluate the trueness and precision of the QuickMIC system, the repeatability and reproducibility between laboratories, as well as to gain insight on performance of the system in real-life microbiological workflows.

## 2 Methods

### Study locations

The data was collected from February to November 2022 at four separate locations; three clinical microbiological laboratories and the internal laboratory at Gradientech. The internal laboratory ran spiked samples and performed the BMD reference testing together with Eurofins Pegasuslab AB, Uppsala, Sweden (about 25% of tests). The three clinical laboratories were located in Uppsala and Örebro, Sweden, as well as Hamburg, Germany (Table 1). At the clinical sites, consecutive Gram-negative samples were run on the QuickMIC system, with the inclusion criteria being Gram-negative with monomicrobial presentation under microscope. Confirmed polymicrobial samples and species not covered by the QuickMIC IFU were excluded from analysis.

**Table 1.** Details of the study locations.

|  | **Gradienttech** | **Örebro University Hospital** | **Uppsala University Hospital** | **University Medical Center Hamburg-Eppendorf** |
| --- | --- | --- | --- | --- |
| **Location** | Uppsala, Sweden | Örebro, Sweden | Uppsala, Sweden | Hamburg, Germany |
| **Size (beds)** | - | Small (460) | Medium (850) | Large (1700) |
| **Blood culture sets/year** | - | 20 000 BCs | 27 000 BCs | 25 000 BCs |
| **Blood culture system** | BACTEC™ FX40 (BD) | BACTEC™ FX (BD) | BACT/ALERT® VIRTUO® (bioMérieux) | BACTEC™ FX (BD) |
| **Bacterial ID method** | MALDI-ToF | MALDI-ToF (from positive BC vial) | MALDI-ToF (from shortly incubated agar plate (4h)) | MALDI-ToF (from shortly incubated agar plate (4h)) |
| **Samples tested** | Challenge, Repeatability | Clinical, Repeatability | Clinical, Repeatability | Clinical, Repeatability |
| **S samples (%)\*** | 37.2 | 80.4 | 75.0 | 70.3 |
| **R samples (%)\*** | 62.8 | 19.6 | 25.0 | 29.7 |
| **MDR samples (%)\*** | 40.9 | 3.9 | 5.0 | 5.4 |

\* S: susceptible to all antibiotics tested, R: resistant to at least one tested antibiotic, MDR: resistant to at least three different classes of antibiotic.

### QuickMIC testing with spiked challenge isolates

Trueness evaluation was performed with a reference collection of 411 bacterial challenge isolates with high resistance rates, acquired from multiple sources (Supplementary Table 1). The isolates were chosen to include a wide variety of susceptibility profiles for each antibiotic on the QuickMIC GN panel. The species distributions are presented in Table 2.

**Table 2.** Species distribution of tested isolates.

|  | *Citrobacter* spp. | *Enterobacter cloacae* complex | *Proteus* spp. | *Pseudomonas aeruginosa* | *Serratia marcescens* | *Klebsiella* spp. | *Escherichia coli* | *Acinetobacter baumannii* complex | Total |
| --- | --- | --- | --- | --- | --- | --- | --- | --- | --- |
| Clinical | 3 | 3 | 4 | 6 | 8 | 28 | 96 | - | 148 |
| Challenge | 15 | 29 | 19 | 58 | 13 | 129 | 128 | 20 | 411 |
| Total | 18 | 32 | 23 | 64 | 21 | 157 | 224 | 20 | 559 |
| Reproducibility | - | 2 | - | 2 | - | 3 | 3 | - | 10 |

The bacterial isolates were streaked on agar plates, grown overnight, and the next day harvested into freezing buffer. The isolates were kept frozen at -70°C for the remainder of the study. All isolates were cultivated using Müller-Hinton (MH) II-agar or MH-II broth (BBL, Becton Dickinson).

Spiked blood cultures for QuickMIC testing were prepared by streaking frozen isolates on plate and incubating overnight at 37°C. The next day, 2-4 colonies were resuspended in MH-II, adjusted to 0.5 McFarland and diluted to a target concentration of 100 cfu/mL into human donor blood or citrated horse blood, after which 10 mL of the blood mixture was used to inoculate blood culture bottles (BACTEC™ PLUS - Aerobic/F Medium, BD), resulting in a start concentration in the blood bottles of 25 cfu/mL. The spiked blood culture bottles were then incubated in a BACTEC™ FX40 system (BD) until a positive signal was received. After positivity, the bottle was removed for QuickMIC testing using the QuickMIC GN cassette (art. nr: 43001). QuickMIC testing was performed as per the protocol of the manufacturer. In brief, ca. 10 μL of positive blood culture (PBC) is added to a sample vial using a standard inoculation loop. The vial contents are filtered to remove cells and debris and injected in the cassette, the gel is allowed to solidify 15 min and afterwards the cassette is loaded into the instrument.

### QuickMIC testing of reproducibility isolates

For measuring the reproducibility of the QuickMIC system, 10 isolates of the reference collection were sent to every location and named RP1 – RP10 (Table 2). The isolates were prepared for QuickMIC using the manufacturer’s protocol for running isolated colonies from plate. In brief, the bacteria were streaked onto an agar plate, incubated overnight at 37°C and resuspended to 0.5 McFarland in PBS, and 10 μL of this solution was used instead of positive blood culture, otherwise using the standard QuickMIC workflow. The ten isolates were run in at least triplicates (on different days) at each study site. Reproducibility was calculated as the total number of results that span a maximum of 3 two-fold dilutions divided by the total number of results. MIC data expressed in a bi-logarithmic scale was used and only antibiotic-bacteria combinations with applicable breakpoints according to EUCAST guidelines (version 13.0, 2023, available at www.eucast.org) were analyzed.

### QuickMIC testing of clinical samples

For patient samples, positive blood cultures were handled as per the routine protocol at the hospital laboratory, and a sample was run on the QuickMIC system using the QuickMIC GN cassette according to the standard protocol of the manufacturer. The samples were further subcultured, identified and handled according to the routine workflow at the clinical laboratory. Bacterial isolates were prepared from the subculture plates and shipped to Gradientech for BMD reference testing. Collected isolates were stored in -70°C during the duration of the study.

### Broth microdilution reference testing

Broth microdilution was performed according to ISO 20776-1:2019 (International Organization for Standardization [ISO]) using pre-filled plates (Merlin Diagnostika GmbH, Bornheim-Hersel, Germany) The antibiotics and concentrations used are described in Supplementary Table 2. In brief, the plates were inoculated using a bacterial suspension at 0.5 McFarland, yielding a final concentration of ∼5*10^5^ CFU/mL per well, and incubated overnight at 37°C. After 18-20h, the MIC was determined according to guidelines in the EUCAST reading guide for broth microdilution v 4.0 (available at www.eucast.org). All isolates were tested at least twice on different days and the modal MIC was determined. If no modal MIC could be achieved after 5 replicate tests, the MIC value was reported as “Not available”. The RP isolates were tested up to 14 times each from different inoculates and by different operators to quantify the variability in the reference BMD-method.

### Data analysis

The QuickMIC AST results, date and time of blood culture start, positivity, and unloading, as well as the start and end times of QuickMIC testing were recorded. QuickMIC MIC results generated by the Analyst software (v 1.0.7) were compared with reference results as previously described [13]. Briefly, linear-scale MIC values are automatically right-censored to nearest log2 dilution step in an equivalent BMD assay, and essential agreement (results differing within 1 log2 step), categorical agreement (results differing in interpreted category), very major (VME), major (ME) and minor error (mE) and bias were calculated as per ISO20776-2:2021. When the reference method showed results below or above the limit of quantification (LOQ) for QuickMIC, these were truncated to the QuickMIC range, as specified by ISO20776-2:2021. The categorization was performed by applying EUCAST clinical breakpoints (version 13.0, 2023, available at www.eucast.org). Linear and logistic regression was performed in R (v 4.3.1) to investigate parameters influencing the result quality. For reproducibility analysis, the mean MIC and standard deviation (SD) of MIC of all runs were calculated, and the SD normalized to the target MIC for each tested QC strain and antibiotic.

### Ethics

The study protocol for the clinically derived samples was reviewed by the Swedish Ethical Review Authority (Dnr: 2020-03060) and by the ethical committee of the chamber of physicians in Hamburg, Germany (Ref: 2021-300120-WF).

## 3 Results

### Reproducibility of QuickMIC between laboratory sites

The overall reproducibility was 98.9% of results within one log2 step between the sites, calculated from the log2 QuickMIC result. In total, 36% of all data points were on scale. The precision of the linear MIC value was also evaluated and calculated as the standard deviation between linear MIC replicates. For the complete dataset, the median standard deviation was 3.0%, and for the included antibiotics ranged between 1.3% and 8.6% per antibiotic (Table 3). The variation in the log2 MIC value was further compared to the variation in the BMD method (Fig. 1, Supplementary fig. 1), indicating that the QuickMIC sample-sample variation was lower than BMD. On average for all antibiotics, 29% vs. 13% of all replicates deviated from the consensus MIC using the BMD method and QuickMIC, respectively, corresponding to an overall 2.2x higher reproducibility (range 1-11x higher, per antibiotic).

**Table 3.** Results from reproducibility and precision testing per antibiotic. The table summarises the reproducibility (number of datapoints within allowable variation as fraction of all datapoints), as well as the precision of the continuous scale linear MIC, as standard deviation (SD) of the MIC for each antibiotic-bacteria target normalized to each maximum concentration tested.

| Antibiotic | Number of data points on scale | Number of data points within ±1 dilution step | Total number of data points | Reproducibility (%) | Median SD of linear MIC (%) |
| --- | --- | --- | --- | --- | --- |
| AMI | 94 | 127 | 127 | 100 | 4.3 |
| CEP | 35 | 128 | 131 | 97.7 | 3.2 |
| CIP | 38 | 132 | 132 | 100 | 3.0 |
| COL | 17 | 120 | 120 | 100 | 1.3 |
| CTA | 25 | 106 | 107 | 99.1 | 5.3 |
| CTV | 38 | 125 | 128 | 97.7 | 3.0 |
| CTZ | 40 | 133 | 133 | 100 | 3.4 |
| GEN | 44 | 105 | 106 | 99.1 | 2.1 |
| MER | 33 | 110 | 115 | 95.7 | 8.6 |
| PIT | 50 | 124 | 124 | 100 | 1.9 |
| TIG | 21 | 38 | 39 | 97.4 | 3.0 |
| TOB | 60 | 99 | 100 | 99 | 5.4 |
| Total | 495 | 1347 | 1362 | 98.9 | 3.0 |

### QuickMIC performance on challenge and clinical isolates

The challenge isolate collection consisted of a selection of common as well as more rarely encountered species in positive blood cultures, with MIC values around the clinical breakpoints to a high degree. All the species included in the study and their respective numbers are depicted in Table 2. The overall essential agreement, categorical agreement and error rates for the challenge isolates are presented in Table 4a. Performance per antibiotic were overall acceptable at >90% EA, CA and bias within ±30%, except for meropenem and piperacillin/tazobactam. The QuickMIC system produced MIC results within 2-4 hours, excluding sample preparation. For the clinical samples, 148 samples were collected from the three locations, representing most species commonly encountered in bloodstream infections. A more detailed breakdown of the susceptibility profiles for each species and each antibiotic is shown in Fig. 2. The time from start of the incubation of the blood culture bottles to QM result (total turnaround time, TTAT) was on average 27.4h for the clinical samples, and the time from blood culture unload to AST result (turnaround time, TAT) was on average 9.2h (SD: 4h) (Supplementary Table 3, Fig. 3a). CA and EA rates were overall >90% for all tested antibiotics compared to BMD. No systematic differences could be seen between challenge and clinical isolates. QuickMIC performance for the complete dataset (clinical and challenge datasets together, Table 4c) was overall good with EA and CA >90% for all tested antibiotics, and average time to result of 3h and 17 min.

**Table 4a.** Results for spiked challenge isolates.

|  | N<sub>results</sub> | CA | EA | S | I | R | mE | ME | VME | Bias | Mean time (h:m) |
| --- | --- | --- | --- | --- | --- | --- | --- | --- | --- | --- | --- |
| AMI | 345 | 333 (96.5) | 315 (91.3) | 297 | 0 | 48 | 0 (0) | 2 (0.6) | 10 (2.9) | 14.73 | 2:34 |
| CEP | 312 | 281 (90.1) | 287 (92) | 173 | 20 | 119 | 26 (8.3) | 1 (0.3) | 4 (1.3) | -20.50 | 3:27 |
| CIP | 378 | 365 (96.6) | 372 (98.4) | 170 | 36 | 172 | 13 (3.4) | 0 (0) | 0 (0) | -7.26 | 3:32 |
| COL | 289 | 283 (97.9) | 279 (96.5) | 232 | 0 | 57 | 0 (0) | 0 (0) | 6 (2.1) | -25.03 | 3:04 |
| CTA | 309 | 305 (98.7) | 307 (99.4) | 159 | 7 | 143 | 4 (1.3) | 0 (0) | 0 (0) | -4.70 | 3:36 |
| CTV | 330 | 324 (98.2) | 320 (97) | 292 | 0 | 38 | 0 (0) | 6 (1.8) | 0 (0) | -15.70 | 3:12 |
| CTZ | 345 | 329 (95.4) | 335 (97.1) | 158 | 45 | 142 | 15 (4.3) | 0 (0) | 1 (0.3) | 4.89 | 3:33 |
| GEN | 313 | 306 (97.8) | 305 (97.4) | 237 | 0 | 76 | 0 (0) | 4 (1.3) | 3 (1) | 19.14 | 3:11 |
| MER | 362 | 315 (87) | 322 (89) | 280 | 25 | 57 | 35 (9.7) | 0 (0) | 12 (3.3) | -40.66 | 3:25 |
| PIT | 317 | 285 (89.9) | 286 (90.2) | 171 | 33 | 113 | 18 (5.7) | 4 (1.3) | 10 (3.2) | -17.83 | 3:30 |
| TIG | 114 | 114 (100) | 112 (98.2) | 113 | 0 | 1 | 0 (0) | 0 (0) | 0 (0) | * | 3:06 |
| TOB | 337 | 332 (98.5) | 329 (97.6) | 227 | 0 | 110 | 0 (0) | 2 (0.6) | 3 (0.9) | 29.84 | 3:07 |
| Overall | 3,751 | 3572 (95.2) | 3569 (95.1) | 2,509 | 166 | 1,076 | 111 (3) | 19 (0.5) | 49 (1.3) | -4.52 | 3:17 |

\* Not evaluable, too few on-scale results

**Table 4B.** Results for clinical isolates.

|  | N<sub>results</sub> | CA | EA | S | I | R | mE | ME | VME | Bias | Mean time (h:m) |
| --- | --- | --- | --- | --- | --- | --- | --- | --- | --- | --- | --- |
| AMI | 138 | 137 (99.3) | 128 (92.8) | 137 | 0 | 1 | 0 (0) | 1 (0.7) | 0 (0) | 22.02 | 2:26 |
| CEP | 142 | 134 (94.4) | 134 (94.4) | 132 | 5 | 5 | 7 (4.9) | 1 (0.7) | 0 (0) | -30.25 | 3:12 |
| CIP | 138 | 129 (93.5) | 135 (97.8) | 114 | 10 | 14 | 8 (5.8) | 1 (0.7) | 0 (0) | -9.72 | 3:14 |
| COL | 127 | 127 (100) | 125 (98.4) | 115 | 0 | 12 | 0 (0) | 0 (0) | 0 (0) | -34.33 | 2:59 |
| CTA | 131 | 130 (99.2) | 130 (99.2) | 122 | 1 | 8 | 0 (0) | 1 (0.8) | 0 (0) | -14.96 | 3:23 |
| CTV | 142 | 141 (99.3) | 140 (98.6) | 142 | 0 | 0 | 0 (0) | 1 (0.7) | 0 (0) | -15.77 | 2:55 |
| CTZ | 143 | 138 (96.5) | 138 (96.5) | 128 | 8 | 7 | 3 (2.1) | 2 (1.4) | 0 (0) | -5.88 | 3:16 |
| GEN | 117 | 114 (97.4) | 111 (94.9) | 112 | 0 | 5 | 0 (0) | 3 (2.6) | 0 (0) | 30.89 | 3:01 |
| MER | 138 | 138 (100) | 137 (99.3) | 138 | 0 | 0 | 0 (0) | 0 (0) | 0 (0) | 2.17 | 3:06 |
| PIT | 134 | 129 (96.3) | 128 (95.5) | 123 | 5 | 6 | 1 (0.7) | 1 (0.7) | 3 (2.2) | -29.95 | 3:15 |
| TIG | 82 | 82 (100) | 79 (96.3) | 82 | 0 | 0 | 0 (0) | 0 (0) | 0 (0) | * | 3:03 |
| TOB | 125 | 123 (98.4) | 118 (94.4) | 120 | 0 | 5 | 0 (0) | 2 (1.6) | 0 (0) | 29.52 | 2:53 |
| Overall | 1,557 | 1522 (97.8) | 1503 (96.5) | 1,465 | 29 | 63 | 19 (1.2) | 13 (0.8) | 3 (0.2) | -14.41 | 3:04 |

\* Not evaluable, too few on-scale results

**Table 4C.** Summary Overall performance results.

|  | AMI | CEP | CIP | COL | CTA | CTV | CTZ | GEN | MER | PIT | TIG | TOB | Overall |
| --- | --- | --- | --- | --- | --- | --- | --- | --- | --- | --- | --- | --- | --- |
| CA (%) | 97.3 | 91.4 | 95.7 | 98.6 | 98.9 | 98.5 | 95.7 | 97.7 | 90.6 | 91.8 | 100 | 98.5 | 96.0 |
| EA (%) | 91.7 | 92.7 | 98.3 | 97.1 | 99.3 | 97.5 | 96.9 | 96.7 | 91.8 | 91.8 | 97.4 | 96.8 | 95.6 |
| mE (%) | 0.0 | 7.3 | 4.1 | 0.0 | 0.9 | 0.0 | 3.7 | 0.0 | 7.0 | 4.2 | 0.0 | 0.0 | 2.4 |
| ME (%) | 0.6 | 0.4 | 0.2 | 0.0 | 0.2 | 1.5 | 0.4 | 1.6 | 0.0 | 1.1 | 0.0 | 0.9 | 0.6 |
| VME (%) | 2.1 | 0.9 | 0.0 | 1.4 | 0.0 | 0.0 | 0.2 | 0.7 | 2.4 | 2.9 | 0.0 | 0.6 | 1.0 |
| Result time (h:m) | 2:32 | 3:23 | 3:27 | 3:03 | 3:32 | 3:07 | 3:28 | 3:08 | 3:20 | 3:26 | 3:05 | 3:03 | 3:13 |

**Figure 2.**
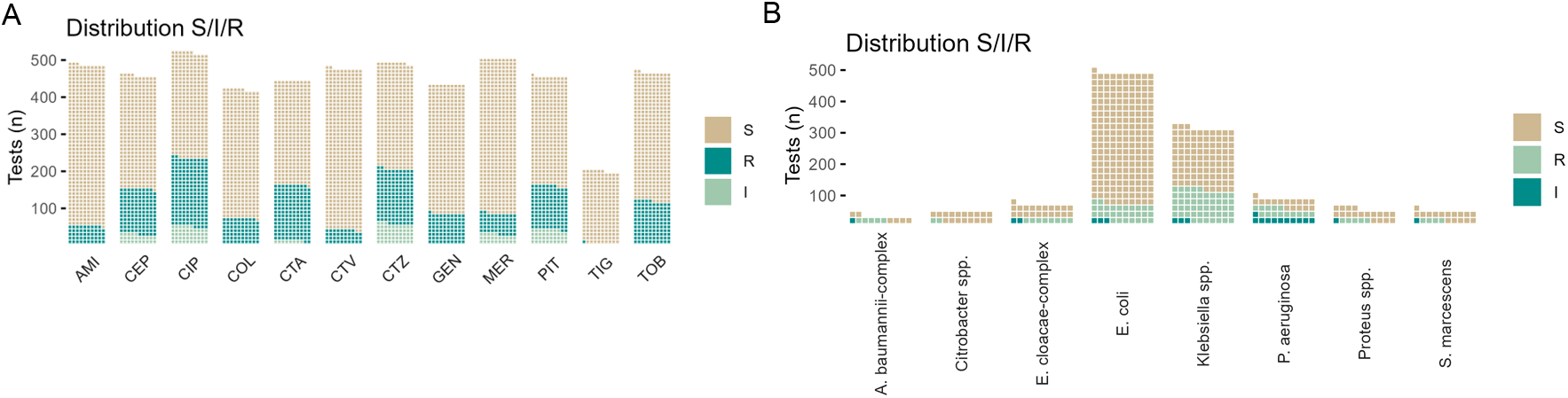
Distribution of susceptibility categories in the complete dataset, split over a) all included antibiotics and b) all included species.

**Figure 3.**
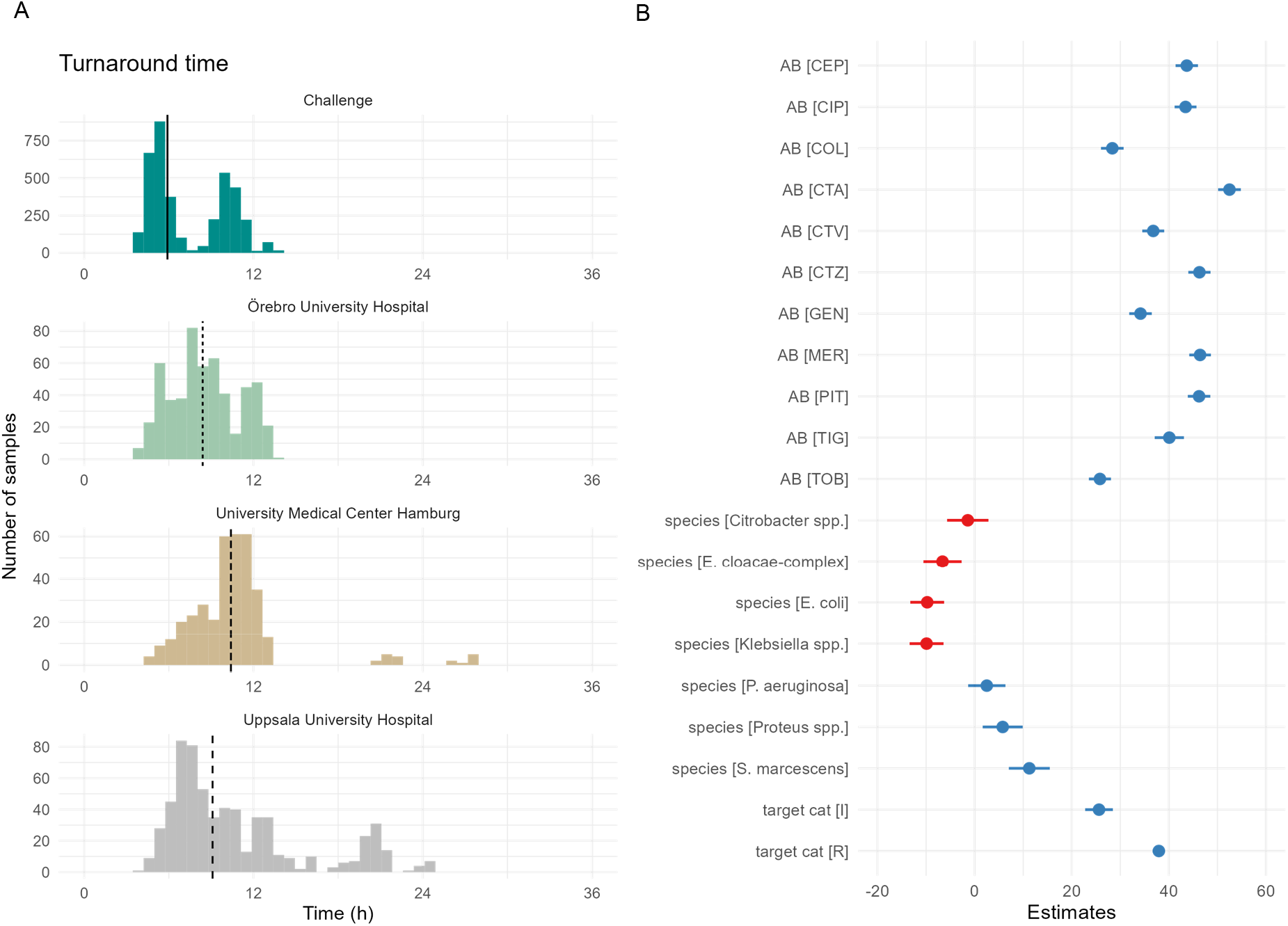
a) Overview of turnaround time (TAT) from the different locations. Lines indicate mean. b) Estimated differences from regression analysis, with contrast set to Antibiotic: AMI, species: Acinetobacter baumannii complex, Target category: S.

### Regression analysis of parameters influencing performance

Regression analysis was used to investigate factors influencing QuickMIC result accuracy and analysis time (Fig. 4). Parameters investigated were reference susceptibility category, species, antibiotics, inoculate concentration of the starting blood culture and delay times from culture bottle positivity and culture bottle unload until start of AST analysis. Resistance category, species and antibiotic were found to significantly affect analysis time, with resistant isolates taking on average 38 minutes longer (p < 0.001) to final result than susceptible ones (Fig. 3b). An overview of the distribution of analysis times over species and antibiotics is displayed in Fig. 4a, b. Regarding effects on accuracy, time from positivity until start of analysis as well as time from unload to start of analysis was not significant in the range from 0-20h (Fig. 4 d, e) (p = 0.16, p = 0.37, respectively). Starting inoculum after the sample preparation step had a significant effect on result accuracy however (p<0.001), with start inoculum concentration <1*10^5^ estimated to provide unacceptable performance (Overall estimated EA<90%, Fig. 4c). Considering this cutoff, 556 of 559 (99.5%) of all tested blood cultures provided an inoculum within acceptable performance range after sample preparation.

**Figure 4.**
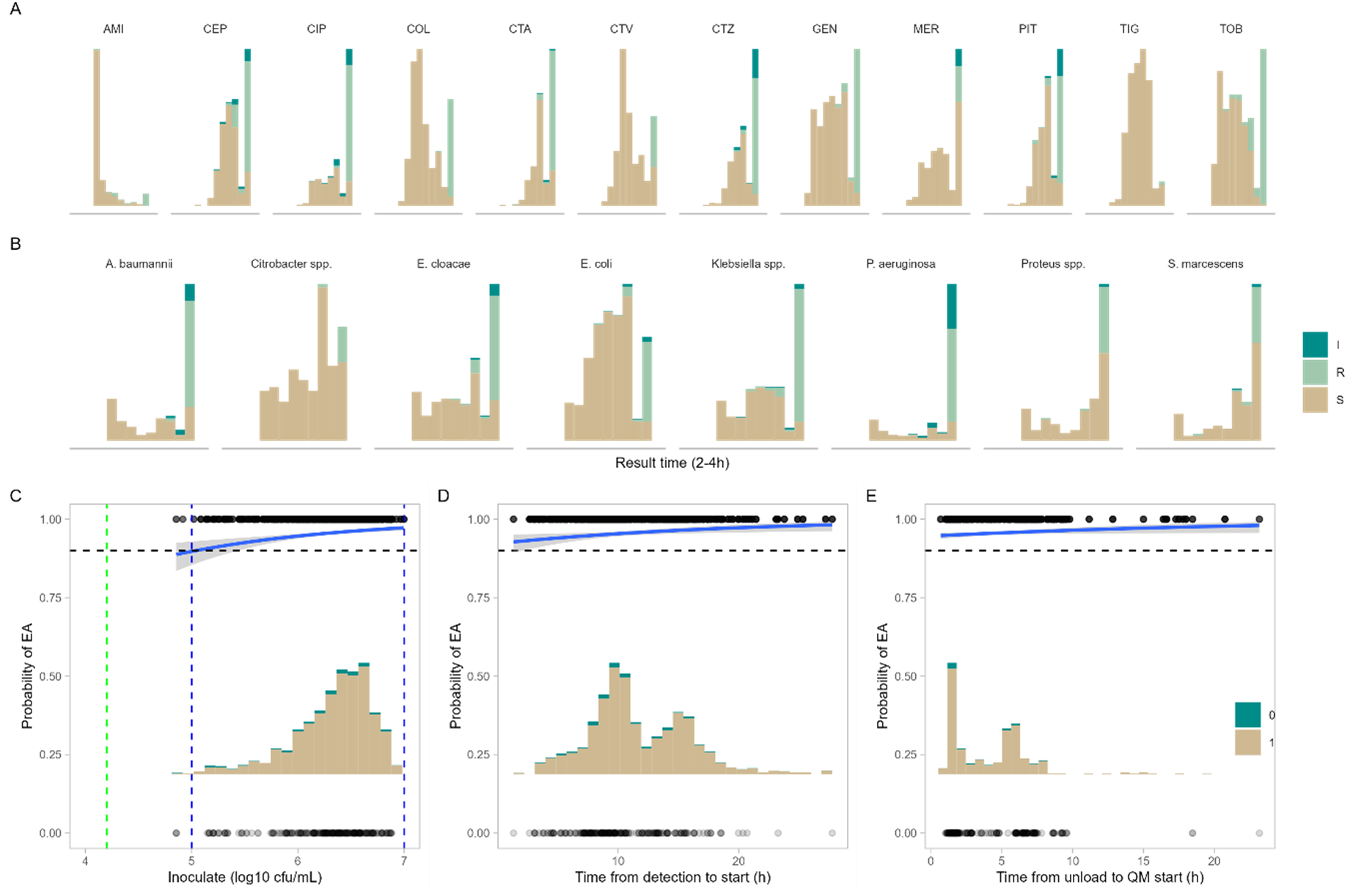
Regression analysis of parameters affecting QuickMIC performance. A and B: analysis time dependence on susceptibility category, where R/I results take longer time than S results, for tested bacteria and antibiotics. C: the ranges of inoculates achieved after sample preparation in comparison to likelihood of achieving accurate results (within EA). Blue lines indicate limits of quantitation, where estimated likelihood of EA>90%, grey field is 95% confidence interval. D and E displays the same analysis for the time from blood culture detection to AST start and time from blood culture bottle unload to AST start, none of which are significantly affecting performance.

## 4 Discussion

The data presented here shows that the QuickMIC rapid AST system has overall good accuracy compared to the reference method broth microdilution, with a significantly improved reproducibility. The three clinical laboratories taking part in the study represented a range of small to large hospital settings and two of the most common blood culture systems in use today, showing wide applicability of the method. The results were of high accuracy also for the challenge dataset, containing a high rate of resistant and multidrug resistant isolates which can be difficult for rapid AST methods to handle due to e.g. delayed resistance [15]. Of note, EA and/or CA for meropenem and piperacillin/tazobactam were lower than 90%, specifically in the challenge dataset, reflecting difficulties. Piperacillin/tazobactam is known to be a problematic antibiotic, for all antibiotic testing methods [16,17]. There are several reasons hypothesized, both relating to bacterial genetics and heteroresistance, as well as stability difficulties with the antibiotic itself. Another issue is relating to the breakpoints, where piperacillin/tazobactam breakpoints lack the I category, and the S/R breakpoint is positioned directly next to the epidemiological cut-off (“wild-type” population). This means that very small errors, even MIC results which are inside essential agreement with reference method, may result in major or very major categorical errors for strains with MIC close to the breakpoint. However, the bias was high for piperacillin/tazobactam (−17.83%) and outside the recommended 30% for meropenem, meaning a systematic undercalling of the meropenem MIC, which will be investigated further. Adjustments to cassette manufacturing and the analysis algorithm may be necessary to improve the bias. Strikingly, 8/12 VME for meropenem were isolates from the CDC AR isolate bank. Further analysis whether specific acquired resistance genes are over-represented in these strains is ongoing, and outside the scope of this study.

Several rAST systems have emerged on the market in recent years in part due to a concerted effort from the public sector involving building awareness as well as economic push and pull incentives [18–20]. Examples include Pheno™ (Accelerate Diagnostics), dRAST™ (Quantamatrix), ASTar® (Q-Linea), Reveal® (Biomerieux, formerly Specific Bioscience) as well as the RAST method from EUCAST. These methods overall display good accuracy >90% and average time to result of 4-7h, which is markedly slower than the 2-4h presented here. The precise, microfluidically generated linear gradient used in QuickMIC is the main differentiator to other rapid AST technologies available today. The exact gradient [13] allows high repeatability and precise MIC results, as demonstrated here. The clinical value of the added precision from a linear MIC, with a linear measurement error, remains to be explored. However, the increased precision also manifests in the reduced variability in the log2 MIC result demonstrated here, which may be beneficial for example in situations where the MIC is close to the clinical breakpoint. Higher repeatability may be beneficial since laboratories often perform single-sample testing due to cost. Greater confidence in MIC values might also lead to novel applications in e.g. precision dosing and personalized medicine in infectious disease, such as PK/PD optimized dosing approaches incorporating the MIC in dosage adjustment [21–23]. MIC based dosage adjustment as a concept has been criticized due to the high inherent variability of existing AST methods [24], a problem which potentially could be rectified by using a more precise AST system. The possibilities opened by a rapid and more precise MIC-method warrant further studies on these themes.

The tradeoff of using a linear gradient is reduced range in comparison to dilution step-based methods. In this dataset, 31.5% of all QuickMIC results were on-scale (59.0% for BMD). The LoQ of QuickMIC is 5% of the maximum measurable concentration, which translates to approximately 5 dilution steps of range, compared to the 8-12 steps used in BMD. In practice, the QuickMIC measurement range is set to include the clinical breakpoints, with either one or two steps above the R breakpoint or below the S breakpoint (Fig. 5). EUCAST emphasizes that measuring MIC in the low breakpoint regions is of limited clinical use [25], which reduces the implications for this limitation in range. Similar arguments can be made for the extreme high resistance range.

**Figure 5.**
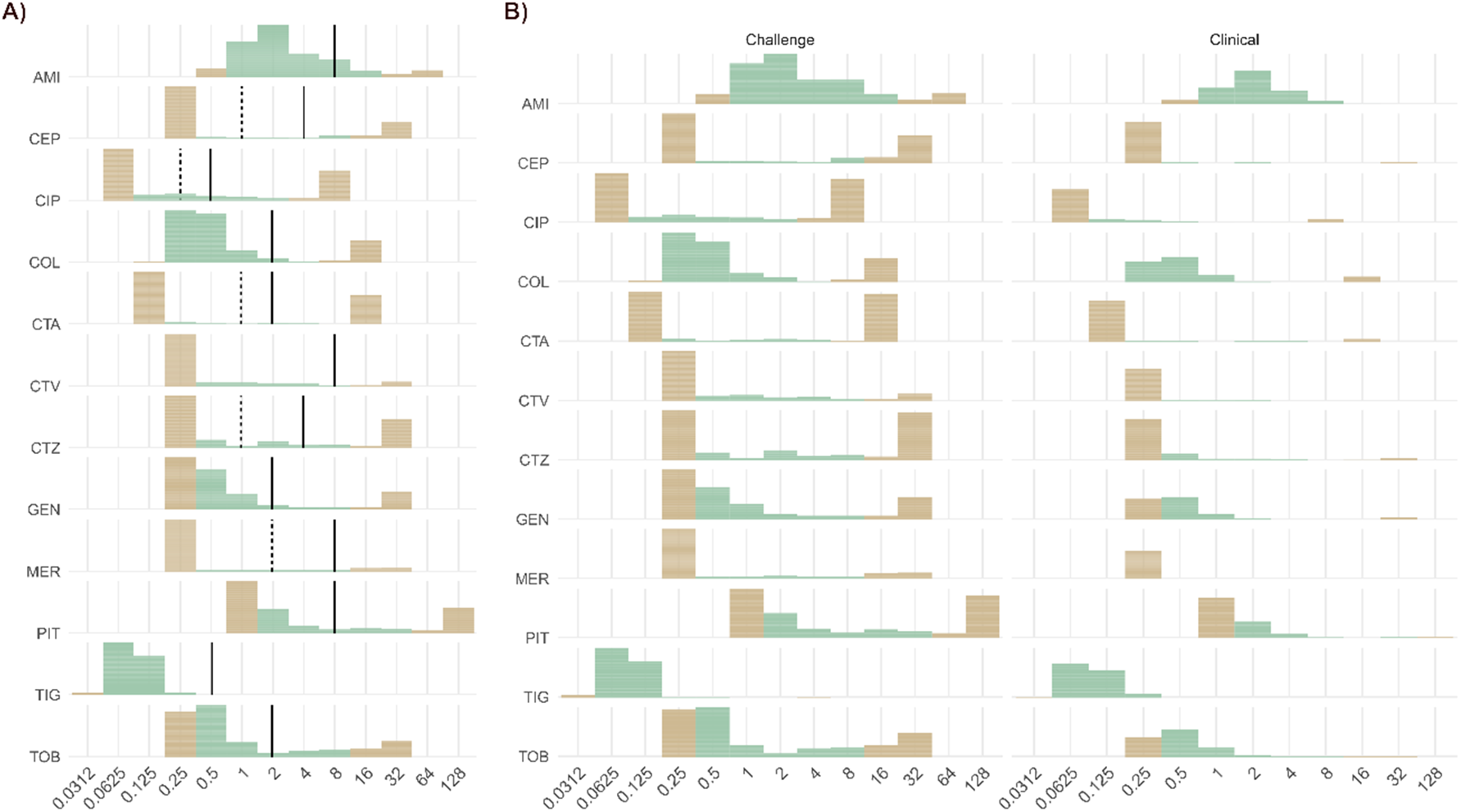
Distribution of MIC results from all strains in study. Color coding according to QuickMIC on-scale or not – green = on QuickMIC scale. Lines indicate I/R breakpoints (I = dashed, R = solid). Right hand side b) shows split by clinical or spiked sample collection.

The inclusion of hospitals only from low to middle resistance setting, in Northern and Central Europe, is a main limitation of this study. It is clear from the performance data that the susceptibility category of the tested isolate has an impact on accuracy and result time. Even though this study tries to counter this bias by inclusion of a highly resistant challenge dataset, convincing data from clinical real-life samples in a high resistance setting is important and needs be investigated further. Other limitations include the handling of polymicrobial samples, which are not supported in QuickMIC and have been excluded from analysis here.

This study demonstrates AST in 2-4 hours directly from positive blood cultures. However, it should be noted that the TAT for the clinical samples in the three locations averaged 9 hours (SD: 4h), which reflects the differences in PBC laboratory workflow. The impact of these procedural differences, manifesting in a delay time from blood culture unloading until AST start ranging from 41min to 23h, were investigated further by regression analysis. While we show that a wide range of delay times from culture positivity as well as from bottle unload to AST start are compatible with accurate rapid AST results, the delay times should be reduced as far as possible for maximum clinical impact of rapid AST.

In conclusion, QuickMIC can be used to rapidly measure MIC directly from blood cultures in clinical settings, with a high reproducibility, precision, and accuracy. The microfluidics-generated linear gradient ensures high repeatability and reproducibility between laboratories, thus allowing a high level of trust in MIC values from single testing, at the cost of reduced measurement range compared to dilution-based methods. The impact on patient outcomes from this technology requires further study however, especially concerning the increased precision of the MIC values produced.

## Data Availability

The datasets presented in this study can be provided on request.

## 6 Conflicts of Interest

ED, JT, LF, JF, AÅ, JB, HÖ, JÅ, CJ and CM are employed by Gradientech AB. TZ was employed by Gradientech throughout the study period (2018-2022). CM owns stock and stock options in Gradientech. The other authors declare no conflict of interest.

## 7 Author Contributions

CM, TZ and CJ planned, funded and supervised the study. BB and CM wrote the manuscript and EG, HR and MS contributed to final review. CM, JÅ, HÖ, TZ and CJ performed data analysis and data management. ED, JT, LF, AÅ, JB, KJ, HA, BB and MR performed the laboratory work.

## 8 Funding

Reagents and consumables were kindly provided by Gradientech AB. The study was partly funded by The Swedish Agency for Economic and Regional Growth (through STUNS, project: BIO-X Accelerate).

## 9 Acknowledgments

The authors thank Eurofins Pegasuslab AB for the contribution to BMD reference testing, and all collaborating laboratories and institutions providing challenge organisms for the study.

## 10 Data Availability Statement

The datasets presented in this study can be provided on request.

## Notes

### Author Declarations

The Swedish Ethical Review Authority waived ethical approval (Dnr: 2020-03060) and the ethical committee of the chamber of physicians in Hamburg, Germany gave ethical approval for this work (Ref: 2021-300120-WF).

### Summary of Updates

Author affiliations updated; funding attribution updated; general spelling updates

