## Supplementary data for "A multicenter evaluation of a novel microfluidic rapid AST assay for Gram-negative bloodstream infections"

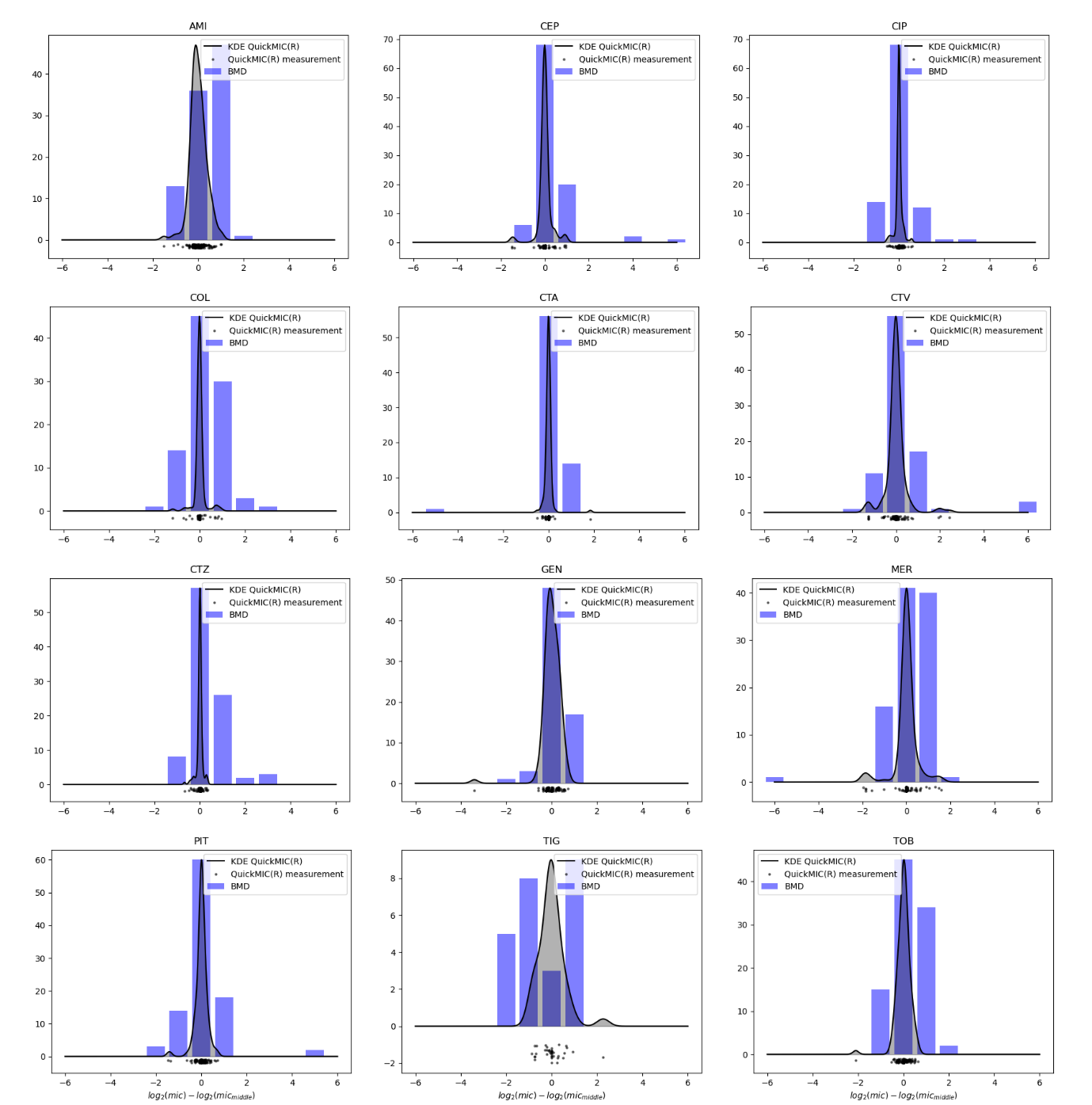

*Supplementary fig. 1 – Comparison between variability from BMD (blue bars) and QuickMIC (dots below the bars, grey shaded field = kernel density estimate for visual comparison of spread, measured by reproducibility isolates run in triplicate at all four study locations (QuickMIC). Overall, QuickMIC results are less variable than BMD for all tested antibiotics.*

*Supplementary table 1 - Origin of challenge isolates:*

Alicante General Hospital, Alicante, Spain;

Antibiotic Research Unit, Uppsala University, Uppsala, Sweden;

The AR ISOLATE BANK, CDC, USA (through the Public Health Agency of Sweden, Solna, Sweden);

The EUCAST Development Laboratory, Växjö, Sweden;

University Clinic of Eppendorf, Hamburg, Germany;

East Tallinn Central Hospital, Tallinn, Estonia;

Uppsala University Hospital, Uppsala, Sweden;

General University Hospital, Valencia, Spain.

*Supplementary table* 2. BMD reference panel

| Antibiotic | | Measuring range (mg/L) |
| --- | --- | --- |
| AMI | Amikacin | 0.5-32 |
| CEP | Cefepime | 0.25-16 |
| CIP | Ciprofloxacin | 0.0625-4 |
| COL | Colistin | 0.125-8 |
| CTA | Cefotaxime | 0.125-8 |
| CTV | Ceftazidime/Avibactam | 0.25-16 |
| CTZ | Ceftazidime | 0.25-16 |
| GEN | Gentamicin | 0.25-16 |
| MER | Meropenem | 0.25-16 |
| PIT | Piperacillin/Tazobactam | 1-64 |
| TIG | Tigecycline | 0.0312-2 |
| TOB | Tobramycin | 0.25-16 |

*Supplementary table 3* – summary of average time performance parameters from the different locations and sample types. Numbers in brackets indicate SD.

|  | BC load to AST end | BC detection to AST end | BC unload to AST end (TAT) | Time in instrument |
| --- | --- | --- | --- | --- |
| Challenge | 23.1 (2.7) | 13.6 (3.1) | 5.9 (2.7) | 3.3 (0.5) |
| Örebro University Hospital | 26.4 (21.2) | 12.6 (6.4) | 8.4 (2.4) | 3.1 (0.4) |
| University Medical Center Hamburg-Eppendorf | 29.9 (26.5) | 16.4 (6.6) | 10.4 (3.7) | 3.1 (0.4) |
| Uppsala University Hospital | 27 (13.9) | 14.5 (5.3) | 9.1 (4.9) | 3 (0.4) |
| Overall clinical | 27.4 (20.3) | 13.8 (6.2) | 9.2 (4) | 3.1 (0.4) |
